# Study on the Intervention of α-Lipoic Acid in Patients with Type 2 Diabetes Mellitus after CAG or PCI Compared with Contrast-Induced Nephropathy

**DOI:** 10.1101/2024.11.15.24317408

**Authors:** Aijie Wu, Chunyao Li, Pingping Wang, Wenlu Shi, Jiaojiao Zhu, Zhengjun Zhang, Dapeng Chen, Ting Zhang

**Affiliations:** Department of Nephrology, General Hospital of Ningxia Medical University, Yinchuan, China; Department of Clinical Medicine, Ningxia Medical University, Yinchuan, China; Department of Cardiovascular Medicine, General Hospital of Ningxia Medical University, Yinchuan, China

**Keywords:** Contrast-induced nephropathy, coronary angiography, percutaneous coronary intervention, α-lipoic acid

## Abstract

**Objective:** To investigate the effect of α-lipoic acid on contrast-induced nephropathy (CIN) in diabetic patients undergoing coronary angiography (CAG) or percutaneous coronary intervention (PCI).

**Methods:** Patients diagnosed with type 2 diabetes mellitus and coronary heart disease scheduled for CAG or PCI treatment at the Department of Cardiovascular Medicine, General Hospital of Ningxia Medical University from February 1, 2021, to August 30, 2023, were recruited. After obtaining informed consent, patients were allocated into three groups: α-lipoic acid group (38 cases), adequate hydration group (60 cases), and routine hydration group (104 cases).The primary outcome observed was the incidence of CIN, and secondary endpoints included changes in SCr, TBiL, and GGT 72 hours after contrast agent administration.

**Results:** The incidence of CIN in the α-lipoic acid group was 2.63% (1/38), 1.67% (1/60) in the adequate hydration group, and 4.81% (5/104) in the routine hydration group, with no statistically significant difference among the three groups (*p*=0.544). After PCI or CAG, SCr levels decreased slightly more in the α-lipoic acid group compared to the adequate hydration group, while the routine hydration group showed an increase, but the differences were not statistically significant (*p*> 0.05).

**Conclusion:** α-Lipoic acid has a certain improvement effect on renal function indicators (Scr) after CAG or PCI, but it did not demonstrate a significant preventive effect on CIN. Adequate hydration showed greater reduction in oxidative stress damage after CAG or PCI compared to α-lipoic acid and routine hydration.

## 1 Introduction

With the increasing use of coronary interventional procedures and radiographic diagnostic techniques, contrast-induced acute kidney injury (CI-AKI) has emerged as the third most common cause^[1]^, in addition to ischemic kidney injury and drug-induced causes ^[1]^. According to the guidelines established by the Kidney Disease International Global Outcomes, contrast-induced nephropathy (CIN) is characterized by a hospitalization-associated increase in serum creatinine (sCr) of 0.3 mg/dL or greater than 1.5 times the baseline value^[2]^. The incidence of CIN in patients undergoing coronary angiography for coronary heart disease is 10% to 15%, but rises to approximately 20% to 40% in patients with concomitant diabetes, mainly related to the kidney damage caused by high blood glucose levels in diabetic patients^[3-6]^. As more evidence accumulates, a declining trend in CI-AKI incidence has been observed. In the United States, the overall incidence of CI-AKI decreased by 26% from 2000 to 2008^[7]^, whether due to the use of different definitions or improved patient management remains unclear. The occurrence of contrast-induced nephropathy is associated with many risk factors, such as hypertension, pre-existing renal impairment, advanced age, and diabetes^[8, 9]^. Pre-existing renal impairment is the strongest risk factor for the development of CIN in patients, and diabetes combined with renal impairment can serve as an independent risk factor for contrast-induced nephropathy. In related studies, hyperglycemia further exacerbates renal function decline, and certain signaling pathways may make diabetes a potential risk factor for contrast-induced renal injury^[10]^. Clinically, diabetic patients are more prone to concomitant coronary artery atherosclerosis, the definitive diagnosis of which relies on coronary angiography (CAG), with some requiring percutaneous coronary intervention (PCI). Once CIN occurs, it prolongs hospital stay, increases patient financial burden, and may even increase mortality rates^[11]^. α-Lipoic acid (ALA), as a B-complex vitamin, is considered one of the strongest natural antioxidants ^[12]^. To date, there have been no clinical studies in China on α-lipoic acid for preventing CIN. This article analyzes whether α-lipoic acid can prevent the occurrence of CIN in diabetic patients, reduce its incidence, and thereby decrease hospitalization time and treatment costs for patients.

## 2 Materials and Methods

### 2.1 General Materials

A total of 202 hospitalized patients with type 2 diabetes mellitus complicated with coronary heart disease, including acute coronary syndrome, stable angina pectoris, and asymptomatic myocardial ischemia, admitted and treated in the Department of Cardiology at General Hospital of Ningxia Medical University from February 1, 2021, to August 30, 2023, were collected. They were scheduled for coronary angiography or coronary stent implantation. Patients were sorted by admission time and divided into three groups: α-lipoic acid (38 cases), adequate hydration group (60 cases), and routine hydration group (104 cases). Among them, the α-lipoic acid group of 38 cases was further divided into CIN group and non-CIN group based on whether CIN occurred in the above three groups.

Inclusion criteria: Hospitalized patients with type 2 diabetes mellitus complicated with coronary heart disease, including acute coronary syndrome, stable angina pectoris, and asymptomatic myocardial ischemia, scheduled for coronary angiography or coronary stent implantation. Exclusion criteria: ➀ Serum creatinine (SCr) > 1.5mg/L (132μmol/L) upon admission; ➁ Malignant tumors; ➂ Liver disease; ➃ Use of nephrotoxic drugs 48 hours before surgery; ➄ Use of other contrast agents one week before surgery; ➅ SCr fluctuation exceeding 5mg/L (442μmol/L) within 48 hours before surgery; ➆ Hypotension, congestive heart failure, or left ventricular ejection fraction <35%; ➇ Regular use of vitamin C for more than one week recently; ➈ Regular use of α-lipoic acid for more than one week recently; ➉ Contrast agent allergy.

### 2.2 Methods

All patients received secondary prevention for coronary heart disease under the guidance of cardiovascular physicians, including enteric-coated aspirin 100mg once daily, ticagrelor 90mg twice daily or clopidogrel hydrogen sulfate 75mg once daily, and statin therapy. The attending physician decided the use of beta-blockers, ACEI/ARB, CCB, diuretics, etc., based on the patient’s condition. All patients underwent laboratory and imaging examinations and signed informed consent forms for surgery. Coronary angiography (CAG) or percutaneous coronary intervention (PCI) was performed via radial or femoral artery puncture by experienced interventional cardiologists. All three groups did not fast before surgery. The routine hydration group received isotonic saline intravenously at a rate of 0.5mL/kg.h for 6-24 hours postoperatively. The adequate hydration group received isotonic saline intravenously at a rate of 1mL/kg.h for 3-12 hours before surgery and 6-24 hours postoperatively, reduced to 0.5mL/kg.h in patients with NYHA class >II heart failure. In addition to routine hydration, the α-lipoic acid group received intravenous infusion of α-lipoic acid injection 0.6g in 250mL 0.9% sodium chloride injection once daily for three consecutive days postoperatively. Based on whether CIN occurred in the above three groups, they were further divided into CIN and non-CIN groups. Standard CAG or PCI procedures were performed, and the contrast agent dose was determined based on patient conditions. Non-ionic low-osmolarity contrast agents iohexol or iopromide and iso-osmolar contrast agent iodixanol were used for all patients, with contrast agent doses recorded intraoperatively.

### 2.3 Observational Indicators

Gender, age, body mass index (BMI), duration of diabetes, surgical method, number of coronary artery lesions, number of stents placed, type and dose of contrast agent, and comorbidities (hypertension, hyperlipidemia). Fasting venous blood samples were collected for Scr, BUN, CysC, TG, IL-6, CRP, BLA, TBiL, HGB, RDW-SD, CTnI, BNP, fasting blood glucose, HbA1c, LVEF; enzyme-linked immunosorbent assay was used to detect oxidative stress markers superoxide dismutase (SOD) and malondialdehyde (MDA). SOD and MDA test kits were purchased from Nanjing Jiancheng Bioengineering Institute, China. Serum creatinine was measured at 72 hours postoperatively. Urine output was recorded, especially within 6 hours postoperatively.

### 2.4 Statistical Analysis

All data were analyzed using SPSS 26.0 software. Measurement data were expressed as mean ± standard deviation (X±S), analyzed using t-test and analysis of variance; count data were expressed as percentages (%), analyzed using Fisher’s exact test. Binary logistic regression analysis was used for risk factor analysis to explore the influence of α-lipoic acid on the risk of CIN occurrence, with odds ratios (OR) and 95% confidence intervals (95%CI) calculated. *p*< 0.05 was considered statistically significant for differences.

## 3 Results

### 3.1 Comparison of Clinical Data Among Three Groups

This study included a total of 218 eligible cases, with 16 cases excluded who did not undergo CAG or PCI (3 in the α-lipoic acid group, 3 in the adequate hydration group, and 10 in the routine hydration group). Ultimately, 202 patients were included in the analysis. Table 1 compares the clinical data of the α-lipoic acid group, adequate hydration group, and routine hydration group. The results indicate that the baseline characteristics among the three groups were similar. There were no statistically significant differences in age, gender, HGB, ALB, TG, LVEF, HbA1c, fasting blood glucose, GGT, TBiL, CTnI, CRP, IL-6, preoperative BUN, preoperative Scr, preoperative SOD activity, preoperative MDA values, duration of diabetes, number of coronary stents implanted, number of coronary artery lesions, type and dose of contrast agent, presence of hypertension, hypertriglyceridemia, use of clopidogrel hydrogen sulfate, enteric-coated aspirin, beta-blockers, ACEI/ARB, CCB, etc. (all *p*> 0.05). Significant statistical differences were observed among the three groups in postoperative SOD, MDA, and surgical methods (*p*< 0.05) (see Table 1). All patients had a urine output between 1000 to 2500 mL per day for three consecutive days before and after surgery, with urine output exceeding 0.5 mL/kg.h.

**Table 1.**
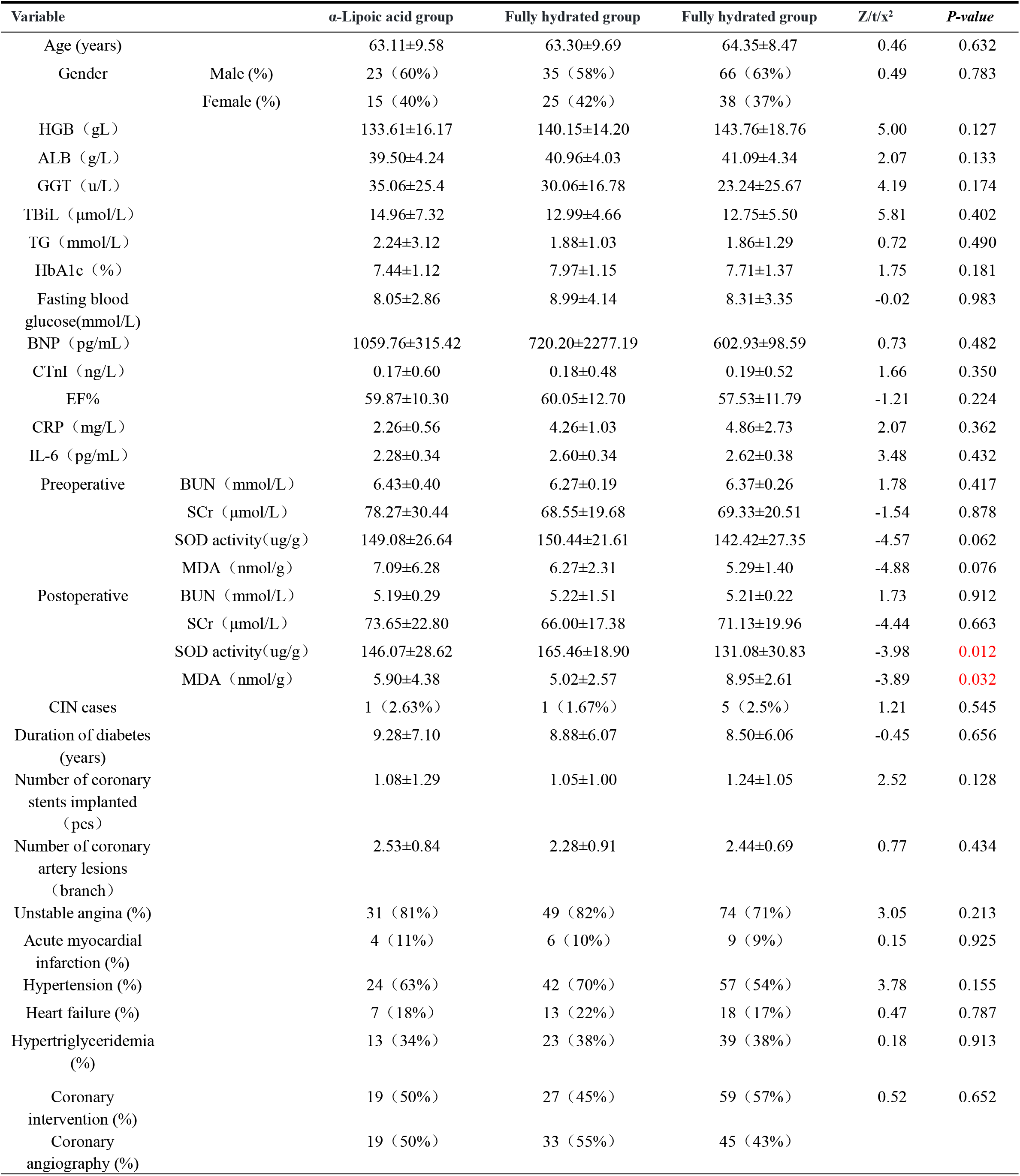
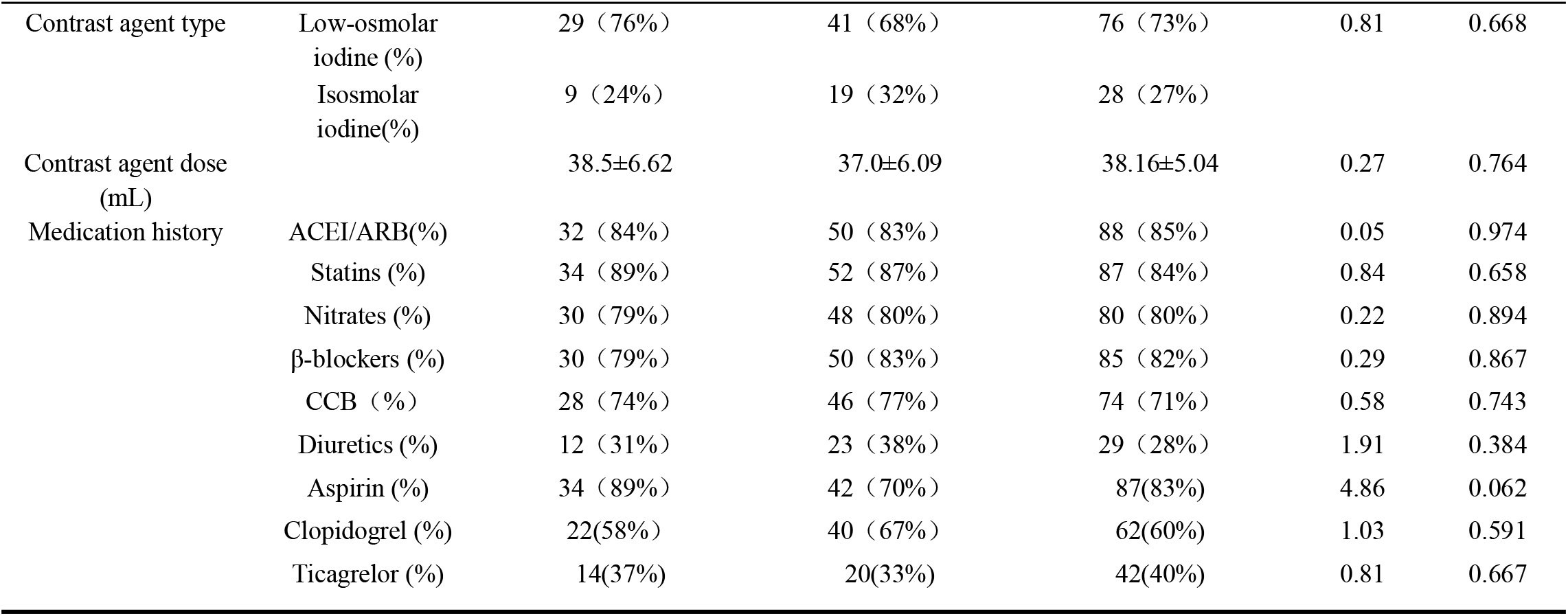
α-lipoic acid group, fully hydrated group, conventional hydration clinical data comparison.

| Variable | | α-Lipoic acid group | Fully hydrated group | Fully hydrated group | Z/t/x<sup>2</sup> | P-value |
| --- | --- | --- | --- | --- | --- | --- |
| Age (years) |  | 63.11±9.58 | 63.30±9.69 | 64.35±8.47 | 0.46 | 0.632 |
| Gender | Male (%) | 23 (60%) | 35 (58%) | 66 (63%) | 0.49 | 0.783 |
|  | Female (%) | 15 (40%) | 25 (42%) | 38 (37%) |  |  |
| HGB (g/L) |  | 133.61±16.17 | 140.15±14.20 | 143.76±18.76 | 5.00 | 0.127 |
| ALB (g/L) |  | 39.50±4.24 | 40.96±4.03 | 41.09±4.34 | 2.07 | 0.133 |
| GGT (u/L) |  | 35.06±25.4 | 30.06±16.78 | 23.24±25.67 | 4.19 | 0.174 |
| TBiL (μmol/L) |  | 14.96±7.32 | 12.99±4.66 | 12.75±5.50 | 5.81 | 0.402 |
| TG (mmol/L) |  | 2.24±3.12 | 1.88±1.03 | 1.86±1.29 | 0.72 | 0.490 |
| HbA1c (%) |  | 7.44±1.12 | 7.97±1.15 | 7.71±1.37 | 1.75 | 0.181 |
| Fasting blood glucose(mmol/L) |  | 8.05±2.86 | 8.99±4.14 | 8.31±3.35 | -0.02 | 0.983 |
| BNP (pg/mL) |  | 1059.76±315.42 | 720.20±2277.19 | 602.93±98.59 | 0.73 | 0.482 |
| CTnI (ng/L) |  | 0.17±0.60 | 0.18±0.48 | 0.19±0.52 | 1.66 | 0.350 |
| EF% |  | 59.87±10.30 | 60.05±12.70 | 57.53±11.79 | -1.21 | 0.224 |
| CRP (mg/L) |  | 2.26±0.56 | 4.26±1.03 | 4.86±2.73 | 2.07 | 0.362 |
| IL-6 (pg/mL) |  | 2.28±0.34 | 2.60±0.34 | 2.62±0.38 | 3.48 | 0.432 |
| Preoperative | BUN (mmol/L) | 6.43±0.40 | 6.27±0.19 | 6.37±0.26 | 1.78 | 0.417 |
|  | SCr (μmol/L) | 78.27±30.44 | 68.55±19.68 | 69.33±20.51 | -1.54 | 0.878 |
|  | SOD activity(ug/g) | 149.08±26.64 | 150.44±21.61 | 142.42±27.35 | -4.57 | 0.062 |
|  | MDA (nmol/g) | 7.09±6.28 | 6.27±2.31 | 5.29±1.40 | -4.88 | 0.076 |
| Postoperative | BUN (mmol/L) | 5.19±0.29 | 5.22±1.51 | 5.21±0.22 | 1.73 | 0.912 |
|  | SCr (μmol/L) | 73.65±22.80 | 66.00±17.38 | 71.13±19.96 | -4.44 | 0.663 |
|  | SOD activity(ug/g) | 146.07±28.62 | 165.46±18.90 | 131.08±30.83 | -3.98 | 0.012 |
|  | MDA (nmol/g) | 5.90±4.38 | 5.02±2.57 | 8.95±2.61 | -3.89 | 0.032 |
| CIN cases |  | 1 (2.63%) | 1 (1.67%) | 5 (2.5%) | 1.21 | 0.545 |
| Duration of diabetes (years) |  | 9.28±7.10 | 8.88±6.07 | 8.50±6.06 | -0.45 | 0.656 |
| Number of coronary stents implanted (pcs) |  | 1.08±1.29 | 1.05±1.00 | 1.24±1.05 | 2.52 | 0.128 |
| Number of coronary artery lesions (branch) |  | 2.53±0.84 | 2.28±0.91 | 2.44±0.69 | 0.77 | 0.434 |
| Unstable angina (%) |  | 31 (81%) | 49 (82%) | 74 (71%) | 3.05 | 0.213 |
| Acute myocardial infarction (%) |  | 4 (11%) | 6 (10%) | 9 (9%) | 0.15 | 0.925 |
| Hypertension (%) |  | 24 (63%) | 42 (70%) | 57 (54%) | 3.78 | 0.155 |
| Heart failure (%) |  | 7 (18%) | 13 (22%) | 18 (17%) | 0.47 | 0.787 |
| Hypertriglyceridemia (%) |  | 13 (34%) | 23 (38%) | 39 (38%) | 0.18 | 0.913 |
| Coronary intervention (%) |  | 19 (50%) | 27 (45%) | 59 (57%) | 0.52 | 0.652 |
| Coronary angiography (%) |  | 19 (50%) | 33 (55%) | 45 (43%) |  |  |

**Table 1(Continued)**
| Variable | | α-Lipoic acid group | Fully hydrated group | Fully hydrated group | Z/t/x<sup>2</sup> | P-value |
| --- | --- | --- | --- | --- | --- | --- |
| Contrast agent type | Low-osmolar iodine (%) | 29 (76%) | 41 (68%) | 76 (73%) | 0.81 | 0.668 |
|  | Isosmolar iodine(%) | 9 (24%) | 19 (32%) | 28 (27%) |  |  |
| Contrast agent dose (mL) |  | 38.5±6.62 | 37.0±6.09 | 38.16±5.04 | 0.27 | 0.764 |
| Medication history | ACEI/ARB(%) | 32 (84%) | 50 (83%) | 88 (85%) | 0.05 | 0.974 |
|  | Statins (%) | 34 (89%) | 52 (87%) | 87 (84%) | 0.84 | 0.658 |
|  | Nitrates (%) | 30 (79%) | 48 (80%) | 80 (80%) | 0.22 | 0.894 |
|  | β-blockers (%) | 30 (79%) | 50 (83%) | 85 (82%) | 0.29 | 0.867 |
|  | CCB (%) | 28 (74%) | 46 (77%) | 74 (71%) | 0.58 | 0.743 |
|  | Diuretics (%) | 12 (31%) | 23 (38%) | 29 (28%) | 1.91 | 0.384 |
|  | Aspirin (%) | 34 (89%) | 42 (70%) | 87(83%) | 4.86 | 0.062 |
|  | Clopidogrel (%) | 22(58%) | 40 (67%) | 62(60%) | 1.03 | 0.591 |
|  | Ticagrelor (%) | 14(37%) | 20(33%) | 42(40%) | 0.81 | 0.667 |

### 3.2 Occurrence of CIN Among Three Groups

The incidence of CIN in the α-lipoic acid group was 2.63% (1/38), in the adequate hydration group was 1.67% (1/60), and in the routine hydration group was 4.81% (5/104). There was no statistically significant difference in the incidence of CIN among the three groups (*p* = 0.685) (see Table 1 and Table 2).

**Table 2.** Comparison of the incidence of CIN in different groups.

| Group | CIN Group (cases) | Non-CIN Group (cases) | Incidence Rate (%) | *P-value* |
| --- | --- | --- | --- | --- |
| α-Lipoic Acid | 1 | 37 | 2.63 | 0.685 |
| Adequate Hydration | 1 | 59 | 1.67 |  |
| Standard Hydration | 5 | 99 | 4.81 |  |

### 3.3 Clinical Data Analysis between CIN and Non-CIN Groups

Patients were divided into non-CIN (195 cases) and CIN groups (7 cases) based on whether CIN occurred post CAG or PCI. The incidence of CIN was 3.47%. Results show no statistically significant differences between the two groups in terms of GGT, TBiL, CRP, preoperative BUN, preoperative Scr, preoperative SOD activity, preoperative MDA values, postoperative BUN, type and dose of contrast agent, etc. (all *p*> 0.05). Significant statistical differences were observed between the two groups in age, postoperative Scr, and postoperative SOD activity (*p* < 0.05) (see Table 3).

**Table 3.** Clinical data analysis of CIN group and non-CIN group.

| Variable |  | Non-CIN Group | CIN Group | t/x<sup>2</sup> | P-value |
| --- | --- | --- | --- | --- | --- |
| Age (years) |  | 63.51±9.01 | 72.14±4.56 | 2.46 | 0.002 |
| GGT (u/L) |  | 27.21±23.45 | 33.20±32.27 | -0.65 | 0.512 |
| TBiL (μmol/L) |  | 14.05±5.76 | 12.67±5.67 | -0.62 | 0.534 |
| CRP (mg/L) |  | 2.16±14.15 | 4.41±2.90 | 0.32 | 0.743 |
| Preoperative | BUN (mmol/L) | 6.36±2.07 | 5.68±2.23 | 0.85 | 0.341 |
|  | Cr (μmol/L) | 70.55±21.47 | 80.15±25.50 | -1.15 | 0.254 |
|  | SOD (ug/g) | 148.87±23.81 | 139.89±11.95 | 1.76 | 0.072 |
|  | MDA (nmol/g) | 7.47±4.63 | 6.02±1.94 | 1.08 | 0.283 |
| Postoperative | BUN (mmol/L) | 5.16±1.64 | 6.27±1.72 | -1.67 | 0.144 |
|  | Cr (μmol/L) | 67.51±17.98 | 109.81±27.19 | -5.99 | 0.015 |
|  | SOD (ug/g) | 137.98±29.42 | 108.68±10.11 | 3.55 | 0.015 |
|  | MDA (nmol/g) | 5.35±3.38 | 8.94±1.74 | -1.03 | 0.022 |
| Contrast agent type | Low-osmolar iodine | 152 (0.78) | 4 (0.57) | 0.69 | 0.406 |
|  | Isosmolar iodine | 43 (0.22) | 3 (0.43) |  |  |
| Contrast agent dose (mL) |  | 38.16±6.99 | 36.81±14.17 | 0.98 | 0.771 |

### 3.4 Risk Factor Analysis for CIN Post CAG or PCI (Logistic Regression Analysis)

CIN was taken as the dependent variable, and potential risk factors influencing CIN were included in a univariate binary logistic regression analysis to further explore factors affecting the risk of CIN occurrence. The results showed that the risk of CIN increased with age (*p*< 0.05). However, variations in ALB, GGT, TBiL, CRP, IL-6, BNP, CTnI, fasting blood glucose, HbA1c, duration of diabetes, number of coronary artery lesions, number of coronary stents implanted, type and dose of contrast agent, and use of α-lipoic acid did not significantly change the risk of CIN occurrence (all *p*> 0.05) (see Table 4).

**Table 4.** Single factor binary Logistic regression analysis results.

| Factor | OR Value | OR Value 95% CI | P-value |
| --- | --- | --- | --- |
| Age | 1.152 | 1.022~1.299 | 0.021 |
| Duration of Diabetes | 0.977 | 0.856~1.114 | 0.729 |
| Number of Coronary Artery Lesions | 0.202 | 0.266~1.323 | 0.593 |
| Number of Coronary Stents Placement | 1.606 | 0.877~2.942 | 0.125 |
| Contrast Agent Dose | 1.632 | 1.054~2.562 | 0.580 |
| Contrast Agent Type | 0.224 | 0.013~3.899 | 0.305 |
| HbA1c | 1.055 | 0.999~1.003 | 0.928 |
| ALB | 1.134 | 0.801~1.606 | 0.479 |
| GGT | 1.035 | 0.953~1.123 | 0.413 |
| TBiL | 1.266 | 0.878~1.400 | 0.207 |
| TG | 0.024 | 0.212~2.011 | 0.099 |
| BNP | 1.001 | 0.999~1.003 | 0.414 |
| CTnI | 1.054 | 0.784~1.485 | 0.764 |
| CRP | 0.891 | 0.585~1.355 | 0.588 |
| ALA | 1.325 | 0.04~38.5 | 0.870 |

### 3.5 Security Assessment

During hospitalization, none of the patients experienced severe adverse events (such as malignant arrhythmia or sudden death). Among patients receiving α-lipoic acid, none reported respiratory distress, rash, itching, taste abnormalities, seizures, diplopia, purpura, platelet dysfunction, acute heart failure, hypotension, etc.,and there were no cases requiring dialysis treatment.

## 4 Discussion

The results of this study show that the overall incidence rate of contrast-induced nephropathy (CIN) was 3.47%. The incidence rates were 2.63% in the α-lipoic acid group (1 case), 1.67% in the adequate hydration group (1 case), and 4.81% in the standard hydration group (5 cases). There was no statistically significant difference in the occurrence of CIN among the three groups (*p*> 0.05). This study selected multiple indicators such as Scr, GGT, and TBiL to comprehensively assess renal function. Although SCr is widely used to detect changes in renal function related to CIN, its levels are affected by skeletal muscle mass and fluid distribution, causing delays in reflecting actual kidney damage. Most patients experience a transient increase in blood creatinine levels 24-48 hours after contrast agent application, with peak values appearing at 3-5 days post-imaging; mild injuries often return to baseline levels within 1-3 weeks^[13]^, indicating that it may not accurately reflect changes in kidney function at an early stage. Moreover, due to the influence of age, sex, and muscle mass, increases in blood creatinine levels often lag behind actual kidney damage, and changes always occur after kidney injury. When the glomerular filtration rate of patients decreases by more than 50%, their blood creatinine levels will show an increase. Nearly 20% of patients cannot even be diagnosed with CIN through blood creatinine levels^[14, 15]^ due to the delay in serum creatinine, so it is necessary to continuously explore more appropriate diagnostic markers. This study preoperatively divided the three groups into Scr: a-lipoic acid group 78.27±30.44 μmol/L, adequate hydration group 68.55±μmol/L, standard hydration group 69.33±20.51μmol/L (*p*= 0.878) no statistical difference; postoperatively three groups Scr: a-lipoic acid group 73.65±22.80μmol/L, adequate hydration group 66.00±17.38 μmol/L, standard hydration group 71.13±19.96 μmol/L (*p*= 0.878) no statistical significance.However, the Scr level in a-lipoic acid group and adequate hydration group decreased slightly compared with that before operation, the decrease in a-lipoic acid group was greater than that in adequate hydration group, and the Scr level in routine hydration group increased slightly compared with that before operation, indicating that α-lipoic acid has a certain improvement effect on renal function after CAG or PCI. These findings provide new theoretical evidence for improving postoperative renal function in patients with type 2 diabetes mellitus and coronary heart disease (CHD) undergoing CAG or PCI.

All patients had a urine output between 1000-2500mL over three days pre- and post-operation, with a urine output greater than 0.5mL/kg.h each day. GGT is widely distributed in various tissues of the body, with the kidneys having the highest concentration, particularly in the proximal tubular cells and biliary system, closely associated with vascular endothelial dysfunction, possibly a crucial factor influencing CIN. In a study by Zheng et al. in 2019^[16]^, they evaluated the relationship between baseline GGT levels and post-PCI heart failure events and found that both decreases and increases in baseline GGT levels increased the incidence of post-PCI heart failure, with a U-shaped curve relationship between GGT and heart failure incidence. This suggests that moderate GGT levels (19.2-39.2u/L) are not conducive to post-PCI heart failure, although the mechanism of this association remains unclear. GGT is believed to be related to inflammation and oxidative stress and has good sensitivity in the early stages of disease. In a study by Oksuz et al.^[17]^, admission GGT was found to be an important and independent predictor of CIN after initial PCI in STEMI patients. Based on these studies, GGT can provide some reference for clinical diagnosis of CIN and prediction of disease outcome. The results of this study show that the preoperative GGT of patients in the CIN group was 33.20±32.27 u/L, higher than the non-CIN group (27.21±23.45 u/L), but the difference was not statistically significant (*p*> 0.05).Total bilirubin (TBiL) includes direct and indirect bilirubin, which are metabolic products of red blood cell aging and breakdown. It is now considered an important endogenous anti-inflammatory and antioxidant molecule that can reduce the formation of peroxide radicals from various biochemical reactions and act as a physiological antioxidant in ischemia^[18]^. In clinical work, it has been found that TBiL levels are significantly lower in patients with necrotizing colitis, intraventricular hemorrhage, and bronchopulmonary dysplasia compared to healthy individuals, indicating that the body consumes TBiL under oxidative stress conditions^[19]^. Higher serum bilirubin levels are associated with lower CIN risk^[20]^. The results of this study show that the preoperative TBiL of patients in the CIN group was 12.67±5.67 μmol/L, lower than the non-CIN group (14.05±5.76 μmol/L), but the difference was not statistically significant.

C-reactive protein (CRP) is an important acute-phase protein, with concentrations increasing over 1000 times in severe inflammatory states^[21]^. Some studies suggest that CRP is an independent predictor of CIN in patients undergoing coronary angiography^[22]^. CRP (C-reactive protein) levels in the CIN group were 4.41±2.90 mg/L compared to 2.16±14.15 mg/L in the non-CIN group, showing higher levels in the CIN group but without statistical significance. Total Superoxide Dismutase (SOD) is a crucial enzyme that helps cells resist oxidative damage, offering protection against oxidative stress and possessing anti-inflammatory and anti-aging properties. Malondialdehyde (MDA) reflects the intensity and rate of lipid peroxidation, indirectly indicating tissue oxidative damage. Preoperatively, there were no statistically significant differences in SOD and MDA levels among the three groups. Postoperatively, SOD levels were significantly different among the groups: α-lipoic acid group (146.07±28.62 ug/g), adequately hydrated group (165.46±18.90 ug/g), and standard hydration group (131.08±30.83 ug/g) (*p*=0.012). This suggests superior antioxidant capacity in the adequately hydrated group. Postoperatively, MDA levels differed significantly among the groups: α-lipoic acid group (5.90±4.38 nmol/g), adequately hydrated group (5.02±2.57 nmol/g), and standard hydration group (8.95±2.61 nmol/g) (*p*=0.032), indicating better cellular protection in the adequately hydrated group.This study reported a CIN incidence of 2.63% in the study group, 1.67% in the adequately hydrated group, and 4.81% in the standard hydration group, with no statistically significant differences (*p*> 0.05). In summary, this study showed that for patients with type 2 diabetes mellitus complicated by coronary heart disease undergoing coronary angiography (CAG) or percutaneous coronary intervention (PCI) treatment, although α-lipoic acid cannot reduce the incidence of CIN, it can slow the postoperative rise in serum creatinine (Scr). This study demonstrated that the adequately hydrated group exhibited the highest postoperative SOD level among the three groups.Thus,the antioxidant effect of the adequately hydrated group was deemed to be more potent than that of the other two groups. Furthermore, the adequately hydrated group displayed the lowest level of MDA among the three groups, suggesting a robust protective effect on cells, the α-lipoic acid group followed closely behind in second place.

Currently, there is no definitive conclusion on the impact of different types of low-osmolar and iso-osmolar contrast agents on the risk of CIN. According to patient height, weight, and renal function, appropriate doses should be selected, generally calculated as 5mL× body weight (kg) / Scr (mg/dl). An ICM dose < 100mL can significantly reduce the incidence of CIN after coronary angiography, with a maximum dose of < 300mL. Therefore, contrast agent dosage is a factor influencing CI-AKI, and in clinical practice, the lowest effective dose should be chosen to meet diagnostic and therapeutic needs ^[2]^. In this study, the α-lipoic acid group received a contrast agent dose of 38.5 ± 6.62mL, the adequately hydrated group received 37.0 ± 6.09mL, and the conventionally hydrated group received 38.16 ± 5.04mL, which are lower doses compared to those reported previously. The contrast agent types were low-osmolar and iso-osmolar, without high-osmolar agents. There were no significant statistical differences in contrast agent types and doses (*p* > 0.05). Moreover, there were no statistically significant differences in contrast agent types and doses between the ICN and non-CIN groups (*p* > 0.05).In this study, the α-lipoic acid group received a contrast agent dose of 38.5±6.62 mL, the adequately hydrated group received 37.0±6.09 mL, and the standard hydration group received 38.16±5.04 ml. These doses were lower compared to previous reports, and the contrast agents used were of low-osmolar and iso-osmolar types, without high-osmolar agents. There were no significant statistical differences observed in the type or dose of contrast agents (*p*> 0.05). Furthermore, there were no statistically significant differences in contrast agent doses or types between the CIN and non-CIN groups (*p*> 0.05).Gender did not affect the incidence of Contrast-Induced Nephropathy (CIN) in this study, whereas age ≥65 years was significantly associated with increased CIN incidence. This age-related increase may be attributed to factors such as age-related decline in renal function, presence of vascular aging, and increased likelihood of other comorbidities exacerbating renal function^[23]^. Regarding age distribution, the α-lipoic acid group had a mean age of 63.11±9.58 years, the adequately hydrated group had 63.30±9.69 years, and the standard hydration group had 64.35±8.47 years, with no statistically significant differences among the three groups. However, the CIN group had a significantly higher mean age of 72.14±4.56 years compared to the non-CIN group (63.51±9.01 years) (*p*=0.002).

This study primarily investigated the impact of α-lipoic acid on the occurrence of Contrast-Induced Nephropathy (CIN) and renal function following coronary angiography (CAG) or percutaneous coronary intervention (PCI). The results indicate that α-lipoic acid did not demonstrate a significant advantage in reducing CIN post CAG or PCI; however, it showed some improvement in postoperative renal function. This provides a theoretical basis for exploring new methods to improve renal function in diabetic patients with coronary artery disease undergoing CAG or PCI. The study showed that adequate hydration reduces the severity of oxidative stress damage. Preoperatively, there were no statistically significant differences in SOD activity and MDA values among the three groups. Postoperatively, the adequately hydrated group exhibited the highest SOD activity, which was statistically significant compared to the other groups (*p*=0.012).Meanwhile, this study has certain limitations. The dose of α-lipoic acid (ALA) used was low, and the treatment duration was short. We administered 600 mg of ALA daily, whereas studies have reported a maximum ALA dose of 1200 mg per day^[24]^. However, the ALA dose used in animal studies was more than 120 times that used in our clinical trial^[25, 26]^. The α-lipoic acid group received standard hydration on top of the protocol, as withholding hydration postoperatively was not feasible and exceeds the recommended usage, which may have affected α-lipoic acid’s effect on postoperative CIN. Further exploration of α-lipoic acid’s effects on CIN post CAG or PCI is warranted in larger, controlled population studies.

This study demonstrated the beneficial effect of α-lipoic acid on postoperative renal function but did not show a significant impact on the occurrence of Contrast-Induced Nephropathy (CIN). There was no statistically significant difference observed (*p*=0.544), which may be attributed to several factors:

1. The study sample size was limited, necessitating larger clinical studies for confirmation.
2. The postoperative observation period was only 72 hours, potentially before serum creatinine (Scr) reached its peak.
3. The duration of medication was short, and the dose was small, possibly not achieving optimal drug concentrations.

The study enrolled patients who received intravenous α-lipoic acid 0.6g daily for three days on top of standard hydration following percutaneous coronary intervention (PCI), without a placebo control group. Postoperative standard hydration may have had some beneficial effect on renal function, potentially masking the effects of α-lipoic acid on postoperative renal function and CIN.Therefore, further evaluation of α-lipoic acid’s preventive effects on CIN post PCI is warranted in larger, controlled studies with diverse populations.

## Data Availability

Data are all available in the article.

## Acknowledgements

The authors thank all the staff of the Department of Nephrology and Cardiovascular Medicine, General Hospital of Ningxia Medical University.

## Statement of Ethics

This study protocol was reviewed and approved by Medical Research Ethics Review Committee of Ningxia Medical University General Hospital, (Approval number: KYLL-2021-371).

